# Increased incidence of thrombosis in a cohort of cancer patients with COVID-19

**DOI:** 10.1101/2020.09.15.20195263

**Authors:** Phaedon D. Zavras, Vikas Mehta, Sanjay Goel, Henny H. Billett

**Affiliations:** Department of Medicine, Jacobi Medical Center, Albert Einstein College of Medicine, Bronx, New York, USA; Department of Otorhinolaryngology, Head & Neck Surgery, Montefiore Medical Center, Albert Einstein College of Medicine, Bronx, New York, USA; Department of Medical Oncology, Montefiore Medical Center, Albert Einstein College of Medicine, Bronx, New York, USA; Division of Hematology, Department of Medical Oncology, Montefiore Medical Center, Albert Einstein College of Medicine, Bronx, New York, USA

**Author notes:** Corresponding Author: Phaedon Dimitrios Zavras, Department of Medicine, Jacobi Medical Center, Albert Einstein College of Medicine, 1400 Pelham Parkway South, Bronx, NY, 10461, USA, /.

**Keywords:** COVID-19, Cancer, Thromboembolic events, Thrombosis

## Abstract

**Background:** Increased rates of TE have been reported in patients with coronavirus disease (COVID-19), even without prior predisposition to thrombosis. Cancer patients are already predisposed to a hypercoagulable state. We aimed to assess whether COVID-19 further increased the risk of thromboembolism (TE) in patients with active cancer.

**Methods:** Data from cancer patients with documented COVID-19 were retrospectively reviewed up to April 10th, 2020. Active cancer was defined as disease treated within the past year. Patients' work-up of thrombosis was done at the clinicians' discretion. All imaging studies' reports within 30 days of the COVID-19 positive test were reviewed for identification of new arterial and/or venous TE. Patients were followed for 30 days from the date of COVID-19+ test for development of TE, hospital length of stay (LOS) and mortality.

**Results:** Of 90 patients, eleven (12.2%) were found to have 13 new TE within 30 days of COVID-19+ test: 8 (8.9%) arterial and 5 (5.6%) venous. Arterial TE were primarily new strokes and/or microvascular cerebral disease (7) with 1 splenic infarct. Venous TE were superficial (1) and deep (3) venous thromboses with 1 pulmonary embolism. Peak D-dimer (DD) values were numerically higher in the TE group vs those with no TE, median peak DD, 7.7 vs 3.2, p=0.25. Out of the patients with CKD/ESRD, 72.7% developed TE vs 31.6% of the those without kidney disease, p=0.02. Hospitalized patients on either prophylactic or therapeutic anticoagulation (AC) had fewer TE than those patients who received no anticoagulation: 9.1% vs 79.0%, p<0.0001. Only 1 patient on enoxaparin prophylaxis developed TE. Mortality was higher in the TE group with a HR of 2.6 for TE vs no TE 95% CI (1.2 - 5.6), p=0.009. Cancer type, disease stage (metastatic vs non), administration of prior chemotherapy, patient setting (inpatient, ICU, outpatient, ED visit), LOS and ventilation did not correlate with increased incidence of TE.

**Conclusion:** Patients with COVID-19 have increased rates of TE, which remained consistent in patients with active cancer within our population. Interestingly, a high incidence of arterial events was noted. TE was associated with worse survival outcomes.

## Introduction

Cancer and the accompanying treatments are well established risk factors for development of arterial (ATE) and, more frequently, venous thromboembolism (VTE) [1]. Cancer increases the risk of thromboembolism (TE) over four-fold that of the general population and, for patients receiving active chemotherapy, the risk is up to 6.5 times greater [2, 3]. For patients actively receiving outpatient chemotherapy, the monthly incidence of VTE has been reported to be 0.8%, which is further increased in those requiring hospitalization [4]. Advanced stages of cancer and the initial period following diagnosis have also been associated with increased TE rates. In addition to the increased morbidity, thromboembolic events are also a leading cause of mortality in this population [5].

Severe acute respiratory syndrome coronavirus 2 (SARS-CoV-2) induced disease (COVID-19) first emerged in December 2020 and was soon declared a pandemic by the World Health Organization (WHO) [6]. As of January 2021, more than 160 million people have been infected worldwide with COVID-19-related mortality reported globally as 2.07% [6]. Although the disease primarily presents with respiratory symptoms, increased thrombosis rates have been observed. A recent meta-analysis reported a 21% VTE rate and a 2% ATE rate in the overall COVID-19 infected population [7]. Patients with COVID-19 and thrombosis have increased mortality over patients without thrombosis [7].

The impact of thrombosis on survival has not, to date, been thoroughly investigated in COVID-19+ cancer patients. A recent retrospective study noted a higher mortality in COVID-19+ patients with active cancer versus a COVID-19+ non-cancer group [8]. Thrombosis rates were increased in both groups relatively to general population, but TE rates were actually lower in the cancer group than in those without cancer: 14.2% (95%CI: 4.7%, 28.7%) vs 18.2% (95% CI, 10.2%, 27.9%) [9]. However, survival was significantly decreased in the cancer group and may have been an interfering risk in this study. There were no data on whether patients with COVID-19 and cancer who developed TE had a worse survival than those who did not develop TE.

Therefore, we sought to quantify the thrombosis events in a real-world study of COVID-19 patients with active cancer during the beginning of the COVID-19 surge at a major tertiary hospital center in New York. Our objectives were to: 1) report the incidence of thrombosis in patients with active cancer and COVID-19; 2) compare the impact of thrombosis on mortality; 3) identify potential risk factors that led to increased thrombosis in this already pre-disposed population, and 4) investigate any potential benefit of pre-emptive anticoagulation.

## Methods

### Study population

A cohort study was conducted in adult patients with active cancer and established COVID-19 infection at Montefiore Healthcare System hospitals who were admitted from March 15, 2020 through April 10, 2020. Active cancer was defined as disease that had been treated within the past year; patients with cancer off treatment for more than 1 year were excluded from the analyses. Demographic, laboratory, radiographic and clinical data were extracted from the electronic medical record (EMR) and hospital databases. The study was reviewed and approved by the Montefiore-Einstein Institutional Review Board (IRB).

### Identification of COVID-19 infection

COVID-19 infection was established with in-house laboratory testing using polymerase chain reaction (PCR) test for SARS CoV-2 from nasopharyngeal swab (RealTime SARS-CoV-2 assay).

### Identification of thrombosis

All imaging studies in the cohort were reviewed for identification of new ATE of VTE. The population of the study was not subject to any imaging hospital protocols. Imaging studies were ordered at the clinicians’ discretion and were solely based on clinical suspicion. Imaging studies were evaluated even if initially ordered for other purposes. Patients with identified atherosclerosis and no TE were classified into the “no thrombosis” group. In the case that an already known TE was re-identified in imaging, it was disregarded. If two studies demonstrated the same TE, this was only counted once. Because shedding of viral particles may have occurred before positive testing and has been reported to still occur as long as 2 months in immunocompromised and cancer patients, we extended the timeframe of our TE identification to 30 days before and 30 days after the initial COVID-19+ test [9]. The reported (if any) d-dimer (DD) values were checked throughout the study period and the peak values were noted.

### Anticoagulation

Prophylactic and therapeutic anticoagulation (AC) data were collected. Institution guidelines recommended that patients admitted with COVID-19 infection and D-dimer (DD) level >3ug/mL or rapid increase of DD be started on therapeutic AC, but the initiation and dosing of AC in the inpatient setting was ultimately at the clinicians’ discretion.

Prophylactic AC was either enoxaparin subcutaneously (SQ) 30mg or 40mg once daily, unfractionated heparin SQ 5000 units two or three times per day or Apixaban 2.5mg twice daily in patients not meeting more than one of the following criteria: age > 80 years, serum creatinine (sCr) ≥1.5mg/dl and weight <60 kilograms (kg). Therapeutic AC was either enoxaparin SQ 1mg/kg twice daily or 1.5mg/kg once daily, continuous heparin infusion (IV), apixaban 5mg (with or without 10 mg loading) or 2.5mg twice daily for patients meeting ≥2 out of the 3 above criteria, rivaroxaban or dabigatran. Patients were classified under the respective groups, i.e. therapeutic AC vs prophylactic AC vs no AC, based on the AC dose administration *before* the development of any new TE.

### Statistical methods

Patients were categorized into two mutually exclusive groups, thrombosis and no thrombosis, based on the identification of new TE within 30 days of COVID+ test. Descriptive statistics were used to summarize demographic and clinical characteristics. Categorical variables were compared using the chi-squared tests and continuous variables were compared using Mann-Whitney rank-sum tests between relevant groups. Kaplan-Meier was used to assess survival between the two groups. Logistic regression was used to define the risk of death. All significance testing was two-sided with a P < 0.05 being indicative of statistical significance. Statistical analyses were performed with IBM SPSS Statistics for Macintosh, Version 27.0. Armonk, NY.

## Results

During the study period, 218 patients with cancer and COVID-19 infection were identified. One hundred and twenty-eight of these patients were excluded because they did not satisfy the active cancer diagnosis. The study cohort consisted of 90 active cancer patients.

Of the 90 patients, 11 (12.2% of the cohort) were found with a new TE during the study period, with two patients having 2 TEs. Eight events (61.5%) were arterial; 7 involved the central nervous system (CNS) with 1 new cerebral vascular accident (CVA) and 6 newly identified microvascular disease, and 1 patient developed new acute splenic infarcts. Five events (38.5%) were venous, including 3 deep venous thrombosis (DVT) events, 1 pulmonary embolism (PE) and 1 superficial venous thrombosis (SVT), which was provoked by a peripheral venous line placement. Out of the 11 patients with new TE, 7 were identified before and 4 after the date of the COVID+ test. All of the new TEs were identified in hospitalized patients; no TEs were identified in the outpatient setting or simple ED visits.

**Table 1** shows the baseline characteristics of the cohort. The median age was 69 years, 62.2% of the population were males and 45.6% were African Americans. Of those with thrombosis, six out of 11 patients with thrombosis (54.5%) were males, 54.5% were African American and 18.2% Hispanic. Sex, race and ethnicity were not different between the groups. Hispanics constituted 31.1% by ethnicity. Common comorbidities were identified between the two groups. Patients with kidney disease had a higher incidence of thrombosis (p=0.02), but this was not true for other comorbidities, such as diabetes mellitus (DM) or obesity. BMI was actually decreased in patients with arterial TE; median BMI (IQR), thrombosis vs no thrombosis, 24.3 (22.3 - 25.9) vs 28.4 (24.9 - 31.8) kg/m^2^, p=0.02.

The incidence of thrombosis was similar for both solid (12.1%) and hematologic (11.4%) malignancies; the distribution of solid or hematologic tumor types was also not associated with thrombosis **(Table 2)**. Importantly, no patients with underlying lung, head and neck, pancreatic, neurologic, hepatobiliary, neuroendocrine malignancies were found to have new thromboses. There was no significant difference in TE between the myeloid and lymphoid malignancy population within the hematologic malignancies. Among the patients with hematologic malignancies, patients with lymphoma, myeloproliferative neoplasms (MPN), acute myeloid leukemia (AML) and chronic lymphocytic leukemia (CLL) did not develop new TE. Metastatic disease, chemotherapy or immunotherapy administration within the past 30 days did not increase the risk of thrombosis.

### Course of illness in patients with COVID-19 and Cancer with and without thrombosis

Table 3 details the course of illness for these patients. Seventy-three patients (81.1% of the cohort) were hospitalized due to COVID-19 at least once during the follow-up period, 8 (8.9%) visited the emergency department (ED) at least once while the remaining 9 patients (10%) were solely followed as outpatients. Of the hospitalized population, 7 patients (7.8% of the cohort, 9.6% of the inpatients) needed intensive care unit (ICU) level of care, whereas 21 (23.3% of the cohort) needed intubation.

D-dimer levels were checked in 45 patients (50% of the cohort) during the study period and 37 patients (41.1% of the cohort) had levels tested during the first 36 hours of admission. The peak D-dimer value, during the total admission time and within 36 hours of admission, was elevated in all patients and was not significantly higher in the thrombosis group **(Table 3)**.

### Anticoagulation data in the inpatient setting

Twelve out of the 73 inpatients (16.4%) received therapeutic AC, 38/73 (52.1%) received prophylactic AC and the rest 23/73 (31.5%) did not receive any form of AC. Four of the 11 patients with thrombosis (36.4%) were identified with their new thromboses on the day of admission, thus were not exposed to any form of prior AC.

Nine patients (75% of patients on therapeutic AC) had been on therapeutic AC at home. Reasons for AC at home included atrial fibrillation (7 patients) and prior VTE (2 patients). The remaining 3 patients were started in-hospital. One of the 3 was initially started on prophylactic AC when admitted but was later switched to therapeutic dose because of concern for PE. Out of the patients who received solely prophylactic dose AC, enoxaparin was given as first agent in 16/38 (42.1%), subcutaneous heparin in 15/38 (39.5%), and 2.5mg dose apixaban in 7/38 (18.4%).

All of the reported TE occurred in hospitalized patients. Of all 11 patients with new confirmed TE, only 1 patient received anticoagulation (prophylactic AC) prior to the thrombotic event. This patient was found to have new microvascular ischemic changes in the cerebral white matter 3 days after being found to be COVID-19+. All other newly confirmed TE (in 10/11 patients, 90.9%) occurred in the setting of no prior AC. Anticoagulation significantly correlated with fewer TE among the inpatients; any AC, TE vs no TE, 9.1% vs 79.0%, p<0.0001 **(Table 3)**.

### Survival

The thrombosis group was found to have significantly decreased survival within the first 30 days following the COVID-19+ test (shown in **Fig. 1**). Univariate analysis showed a hazard ratio (HR) of 2.6 (95% CI:1.2 – 5.6, p=0.0009) for mortality in patients with thrombosis vs no thrombosis. Nine out of 11 patients of the thrombosis group (81.8%) and 33 out of the 79 patients without thrombosis (41.7%) expired. Of note, 7 patients with thrombosis (63.6% of the thrombosis group) expired within the first seven days since COVID-19 diagnosis.

**Fig. 1.**
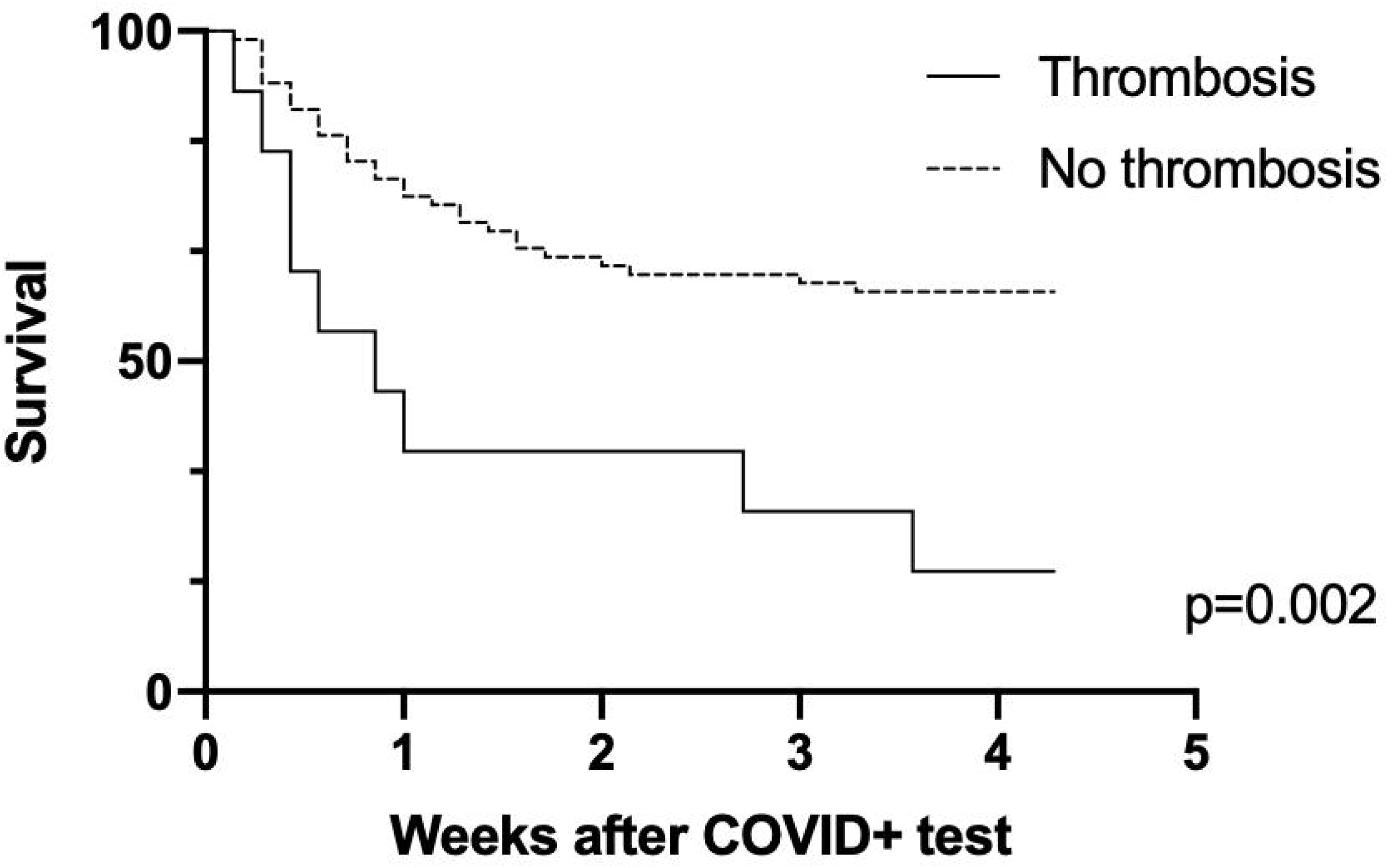
Kaplan-Meier curve of survival between the thrombosis and no thrombosis groups

## Discussion

Cancer is a well-known independent risk factor for thromboembolism and COVID-19 infection has also been associated with a hypercoagulable state. A recent meta-analysis of 8271 COVID-19+ patients revealed 21% VTE and 2% ATE rate [7]. While recent studies have shown that cancer is an independent risk factor for TE in patients with COVID-19 infection [10], the characteristics and impact on survival of patients with COVID-19 and active cancer have not been well described. Whether these two risks are synergistic or additive is unclear but with hallmarks of disease being high fibrinogen and D-dimer levels in addition to elevated inflammatory markers, such as C-reactive protein (CRP), ferritin and interleukin-6 (IL-6) [11], it may be important to determine.

The analysis of the present cohort is in alignment with the recent reports, showing increased monthly incidence of TE at 12.2%, as compared to 0.8% of monthly TE incidence in patients with active cancer and no COVID-19 infection [2]. Interestingly, the majority of our events were arterial (8/13, 61.5%). This can be partly explained by the high number of computed tomography (CT) scans of the head done in this cohort (total 20 CTs), which identified 7/8 ATEs, as compared to studies done to detect VTE (7 CT chest with PE protocol and 12 DVT studies). It is also notable that patients with cancers historically associated with increased thrombosis risk, such as pancreatic cancer and several hematologic malignancies, did not develop any TE. Although many comorbidities, such as diabetes and obesity, have been shown to independently increase the mortality risk in the COVID-19+ population [12, 13], these did not seem to further increase the risk of thrombosis. Kidney disease was significantly associated with increased TE risk, with 8/11 patients with TE (72.7%) having underlying CKD or ESRD.

All of the patients with COVID and TE were inpatients. This is not surprising, given the fact that inpatients were more likely to be afflicted with a severe inflammatory response, which would further exacerbate their prothrombotic state. Mean hospital LOS was numerically higher in the thrombosis group. Admission to the ICU or need of ventilation support was not associated with a higher risk of thrombosis in our cohort.

Prior studies have shown that D-dimer levels seem to correlate well with the risk of thrombosis in COVID-19 infected patients [14]. Patients with active cancer would have been expected to have higher baseline D-dimer and may indicate a further prothrombotic risk in the cancer population. In the early days of the pandemic, our institution had implemented a D-dimer cut-off value of 3ug/mL or rapidly increasing DD in order to initiate empiric therapeutic rather than prophylactic AC. However, our numbers were too small to determine whether this had any effect in our cancer population. Our data suggest that any type of AC, either prophylactic or empiric therapeutic, seemed to protect against TE in our cohort.

## Limitations

The present patient population was characterized by very high mortality following the COVID-19+ test; in total, 29/90 patients (32.2%) expired within one week of COVID-19 diagnosis, 7 from the thrombosis group (63.3% of the group) and 22 from the non-thrombosis group (27.8% of the non-TE group). Since almost one third of the non-thrombosis group evaluated for new TE was censored early since they expired early after diagnosis, the competing mortality risk may have led to underreporting of the real TE risk. Finally, because of the high risk of exposure at the time, thromboembolic events may have been underreported due to the inability to pursue the appropriate imaging tests.

## Conclusion

Our study highlights that thromboembolism imposes an independent risk of mortality in cancer patients with COVID-19. Further studies are needed to confirm our results and further investigate the overall impact of thromboembolism in this patient population.

## Data Availability

The data were extracted from Montefiore Healthcare System EMR.

## Statements

### Statement of Ethics

The present study is in accordance with the World Medical Association Declaration of Helsinki. The Montefiore-Einstein Institutional Review Board (IRB) reviewed and approved the study and waived the need for informed consent.

### Author Contributions

PDZ was responsible for the study design, data collection, data analysis, interpretation of the results, and drafted the initial manuscript. HHB was responsible for the study design, major revisions and final approval of the version submitted. RK, VM and SG were responsible for data collection and revision of the manuscript.

### Data Availability Statement

The data for the present study were extracted from the Montefiore Health Care System’s EMR, thus they are not publicly available because this would compromise the privacy of research participants. Research datasets would be available from PDZ upon reasonable request.

