## Supplementary material for "Increased incidence of thrombosis in a cohort of cancer patients with COVID-19": Table 1

|  | All patients | Thrombosis | No Thrombosis | p |
| --- | --- | --- | --- | --- |
|  | 90 (100.0%) | 11 (12.2%) | 79 (87.8%) |  |
| Age, Median (IQR) | 69 (60 - 78) | 60 (54 - 76) | 71 (61 - 79) | 0.76 |
| Female (N, %) | 34 (37.8%) | 5 (45.5%) | 29 (36.7%) | 0.74 |
| Race | | | | |
| White (N, %) | 10 (11.1%) | 0 (0.0%) | 10 (12.7%) | 0.23 |
| African American (N, %) | 41 (45.6%) | 6 (54.5%) | 35 (44.3%) |  |
| Asian (N, %) | 5 (5.6%) | 2 (18.2%) | 3 (3.8%) |  |
| Other/NA (N, %) | 34 (37.8%) | 3 (27.3%) | 31 (39.2%) |  |
| Ethnicity | | | | |
| Ηispanic (N, %) | 28 (31.1%) | 2 (18.2%) | 26 (32.9%) | 0.49 |
| Νon-Hispanic (N, %) | 58 (64.4%) | 8 (72.7%) | 50 (63.3%) |  |
| N/A (N, %) | 4 (4.4%) | 1 (9.1%) | 3 (3.8%) |  |
| Comorbidities | | | | |
| Diabetes Mellitus (N, %) | 36 (40.0%) | 5 (45.5%) | 31 (39.2%) | 0.75 |
| Hypertension (N, %) | 64 (71.1%) | 9 (81.8%) | 55 (69.6%) | 0.50 |
| Chronic lung disease (N, %) | 23 (25.6%) | 3 (27.3%) | 20 (25.3%) | 0.99 |
| Kidney disease (N, %) | 33 (36.7%) | 8 (72.7%) | 25 (31.6%) | **0.02** |
| Coronary artery disease (N, %) | 13 (14.4%) | 0 (0.0%) | 13 (16.5%) | 0.35 |
| Congestive heart failure (N, %) | 14 (15.6%) | 2 (18.2%) | 12 (15.2%) | 0.68 |
| Obesity (N, %) | 32 (35.6%) | 3 (27.3%) | 29 (36.7%) | 0.74 |

Table 1. Baseline Characteristics of the Study Population
