## Supplementary material for "Increased incidence of thrombosis in a cohort of cancer patients with COVID-19": Table 2

Table 2. Cancer Types in the Study Population

|  | All patients | Thrombosis | No thrombosis | p |
| --- | --- | --- | --- | --- |
|  | N=90 | N=11 | N=79 |  |
| Solid tumors | 58 (100.0%) | 7 (12.1%) | 51 (87.9%) |  |
| Genitourinary | 17 (100.0%) | 2 (11.8%) | 15 (88.2%) | 0.34 |
| Breast | 6 (100.0%) | 1 (16.7%) | 5 (83.3%) |  |
| Colorectal | 6 (100.0%) | 1 (16.7%) | 5 (83.3%) |  |
| Upper gastrointestinal tract | 4 (100.0%) | 1 (25.0%) | 3 (75.0%) |  |
| Pancreas | 2 (100.0%) | 0 (0.0%) | 2 (100.0%) |  |
| Neuroendocrine | 2 (100.0%) | 0 (0.0%) | 2 (100.0%) |  |
| Gynecologic | 3 (100.0%) | 1 (33.3%) | 2 (66.7%) |  |
| Lung | 8 (100.0%) | 0 (0.0%) | 8 (100.0%) |  |
| Head and neck | 4 (100.0%) | 0 (0.0%) | 4 (100.0%) |  |
| Neurologic | 1 (100.0%) | 0 (0.0%) | 1 (100.0%) |  |
| Hepatobiliary | 4 (100.0%) | 0 (0.0%) | 4 (100.0%) |  |
| Skin | 1 (100.0%) | 1 (100.0%) | 0 (0.0%) |  |
| Hematologic malignancies | 32 (100.0%) | 4 (11.4%) | 28 (88.6%) |  |
| Myeloid malignancy | 6 (100.0%) | 1 (16.7%) | 5 (83.3%) | 0.55 |
| Myelodysplastic syndromes | 3 (100.0%) | 1 (33.3%) | 2 (66.7%) |  |
| Myeloproliferative neoplasms | 2 (100.0%) | 0 (0.0%) | 2 (100.0%) |  |
| Acute myeloid leukemia | 1 (100.0%) | 0 (0.0%) | 1 (100.0%) |  |
| Lymphoid malignancy | 26 (100.0%) | 3 (11.5%) | 23 (88.5%) | 0.99 |
| Non-Hodgkin's lymphoma | 8 (100.0%) | 0 (0.0%) | 8 (100.0%) |  |
| Hodgkin's lymphoma | 3 (100.0%) | 0 (0.0%) | 3 (100.0%) |  |
| Chronic lymphocytic leukemia | 2 (100.0%) | 0 (0.0%) | 2 (100.0%) |  |
| Multiple Myeloma | 10 (100.0%) | 2 (20.0%) | 8 (80.0%) |  |
| Acute lymphocytic leukemia | 3 (100.0%) | 1 (33.3%) | 2 (66.7%) |  |
| Metastatic cancer (solid tumors only) | 36 (100.0%) | 4 (11.1%) | 32 (88.9%) | 0.99 |
| Chemotherapy ≤30 days | 36 (100.0%) | 3 (8.3%) | 33 (91.7%) | 0.52 |
| Immunotherapy ≤30 days | 5 (100.0%) | 0 (0.0%) | 5 (100.0%) | 0.99 |
