## Supplementary material for "Increased incidence of thrombosis in a cohort of cancer patients with COVID-19": Table 3

Table 3. Course of illness in patients with COVID-19 and Cancer with and without thrombosis

|  | All patients | Thrombosis | No Thrombosis | p |
| --- | --- | --- | --- | --- |
|  | N=90 (100.0%) | N=11 (100.0%) | N=79 (100.0%) |  |
| Patient setting |  |  |  |  |
| ED only, N (%) | 8 (8.9%) | 0 (0.0%) | 8 (10.1%) | 0.23 |
| Outpatient, N (%) | 9 (10.0%) | 0 (0.0%) | 9 (11.4%) |  |
| Inpatient, N (%) | 73 (81.1%) | 11 (100.0%) | 62 (78.5%) |  |
| Admission days, median (IQR) | 5 (3 - 10.5) | 4 (2 - 19) | 5 (3 - 10) | 0.89 |
| ICU, N (%) | 7 (7.8%) | 1 (9.1%) | 6 (7.6%) | 0.99 |
| Mechanical ventilation, N (%) | 21 (23.3%) | 4 (36.4%) | 17 (21.5%) | 0.28 |
| D-Dimer values |  |  |  |  |
| D-Dimer performed, Ν (%) | 45 (50.0%) | 7 (63.6%) | 38 (48.1%) |  |
| Peak, median (IQR) | 3.6 (1.6 - 9.2) | 7.7 (3.5 - 8.7) | 3.2 (1.4 - 10.3) | 0.25 |
| D-Dimer performed ≤36 hrs, Ν (%) | 37 (41.1%) | 4 (36.3%) | 33 (41.8%) |  |
| Peak, median (IQR) | 2.4 (1.6 - 6.1) | 5.6 (2.4 - 8.5) | 2.4 (1.1 - 5.5) | 0.33 |
| Inpatient Anticoagulation Use |  |  |  |  |
| No inpatient AC, N (%) | 23 (31.5%) | 10 (90.9%) | 13 (21.0%) | **<0.0001** |
| Any inpatient AC, N (%) | 50 (68.5%) | 1 (9.1%) | 49 (79.0%) |  |
| Therapeutic AC, N (%) | 12 (16.4%) | 0 (0.0%) | 12 (19.3%) |  |
| Prophylactic AC, N (%) | 38 (52.1%) | 1 (9.1%) | 37 (59.7%) |  |

Abbreviations: ED, emergency department; IQR, interquartile range; ICU, intensive care unit; AC, anticoagulation
